# Social media discourse and internet search queries on cannabis as a medicine: A systematic scoping review

**DOI:** 10.1101/2022.05.16.22275171

**Authors:** Christine Mary Hallinan, Sedigheh Khademi Habibabadi, Mike Conway, Yvonne Ann Bonomo

## Abstract

The use of cannabis for medicinal purposes has increased globally over the past decade since patient access to medicinal cannabis has been legislated. Yet, evidence of cannabis efficacy for a suite of conditions is only just emerging. Although there is considerable engagement from many stakeholders to add to the evidence base through randomized control trials, many gaps in the literature remain. Data from real-world and patient reported sources can provide opportunities to address this evidence deficit. This real-world data can be captured from a variety of sources such as found in routinely collected health care and health services records that include but are not limited to patient generated data from medical, administrative and claims data, patient reported data from surveys, wearable trackers, patient registries, and social media. In this systematic scoping review, we seek to understand the utility of online user generated text into the use of cannabis as a medicine. The objective of this scoping review is to synthesize primary research that uses social media discourse and internet search engine queries to answer the following questions: (i) Does online user-generated text provide a useful data source for studying cannabis as a medicine? (ii) What are the aims, data sources, and research themes of studies using online user-generated text to discuss the medicinal use of cannabis? For this scoping review we used a framework for systematic reviews and meta-analyses, the PRISMA guidelines to inform our methods. We conducted a manual search of primary research studies which used online user-generated text as a data source using the MEDLINE, Embase, Web of Science, and Scopus databases in October 2022. Editorials, letters, commentaries, surveys, protocols, and book chapters were excluded from the review. Forty-two studies were included in this review, 22 studies used manually labelled data, four studies used existing meta-data (Google trends/geo-location data), two studies used data that was manually coded using crowdsourcing services, and two used automated coding supplied by a social media analytics company, 15 used computational methods for annotating data. Our review reflects a growing interest in the use of user-generated content for public health surveillance. It also demonstrates the need for the development of a systematic approach for evaluating the quality of social media studies and highlights the utility of automatic processing and computational methods (machine learning technologies) for large social media datasets. This systematic scoping review has shown that user-generated content as a data source for studying cannabis as a medicine provides another means to understand how cannabis is perceived and used in the community. As such, it provides another potential ‘tool’ with which to engage in pharmacovigilance of, not only cannabis as a medicine, but also other novel therapeutics as they enter the market.

## Introduction

Genetic analysis of ancients cannabis indicates the plant cannabis sativa was first cultivated for use as a as a medicinal agent up to 2400 years ago (1). From the 1800’s, people in the United States (US), widely used cannabis as a medicine by either prescription or as an over the counter therapeutic (2). Yet by the mid-20th century, cannabis use was prohibited in many parts of the developed world with the passing of legislation in the US, the United Kingdom (UK) and various European countries that proscribed its use (3-6). Since the 2000s, the use of cannabis for medicinal purposes has been decriminalized in many countries including Israel, Canada, Netherlands, Uniter States, United Kingdom, and Australia (7-9). More recently, new evidence regarding the clinical efficacy of cannabis for some conditions (10) has triggered public interest in cannabis and cannabis-derived products (11, 12), resulting in a global trend towards public acceptance, and subsequent legalization of cannabis for both medicinal and recreational use. There is emerging evidence of cannabis efficacy for childhood epilepsy, spasticity, and neuropathic pain in multiple sclerosis, acquired immunodeficiency syndrome (AIDS) wasting syndrome, and cancer chemotherapy-induced nausea and vomiting (13-15). Although researchers are investigating cannabis for treating cancer, psychiatric disorders (16), sleep disorders (17), chronic pain (18) and inflammatory conditions such as rheumatoid arthritis (19), there is currently insufficient evidence to support its clinical use. Scientific studies on emerging therapeutics typically exclude vulnerable populations such as pregnant women, young people, the elderly, patients with multimorbidity and polypharmacy, and this limits the availability of evidence for cannabis effectiveness across these population groups (20).

Cannabis as medicine is associated with a rapidly expanding industry (21). Patient demand is increasing, as is reflected in an increasing number of approvals for prescriptions over time (22), with one study showing that 61% of Australian GPs surveyed reported one or more patient enquiries regarding medical cannabis (23). With this increasing demand, is sophisticated marketing by medicinal cannabis companies that leverages evidence from a small number of studies to promote their products (24, 25). In light of this, concerns regarding patient safety is warranted especially when marketing for some cannabinoid products is associated with inadequate labelling and/or inappropriate dosage recommendations (26). These concerns are compounded by the downscheduling of over-the-counter cannabis products which do not require a prescription (27) and the illicit drug market (28). Given this dynamic interplay between marketing, product innovation, regulation, and consumer demand, innovative methods are required to augment existing established approaches to the surveillance and monitoring of emerging and unapproved drugs.

Although there is considerable engagement from many stakeholders to improve the scientific evidence regarding the efficacy and safety of cannabis through randomized control trials, many gaps remain in the literature (29). Yet data from real-world and patient reported data sources could provide opportunities to address this evidence deficit (30). This real-world data can be captured from a variety of sources such as found in routinely collected health care and health services records that include but are not limited to patient generated data from medical, administrative and claims data, as well as patient reported data from surveys, wearable trackers, patient registries, and social media (31-33).

People readily consult the internet when looking for and sharing health information (34, 35). According to 2017 survey of Health Information National Trends, almost 78% of US adults used online searches first to inquire about health or medical information (34). Data resulting from these online activities is labelled ‘user generated’ and is increasingly becoming a component of surveillance systems in the health data domain (36). Monitoring user-generated data on the web can be a timely and inexpensive way to generated population-level insights (37). The collective experiences and opinions shared online are an easily accessible wide-ranging data source for tracking emerging trends – which might be unavailable or less noticeable by other surveillance systems.

The objective of this systematic scoping review is to understand the utility of online user generated text in providing insight into the use of cannabis as a medicine. In this review, we aim to systematically review existing work that utilizes user-generated content to explore cannabis as a medicine. The objective of this review is to synthesise primary research that uses social media discourse and internet search engine queries to answer the following questions:

- Does online user-generated text provide a useful data source for studying cannabis as a medicine?
- What are the aims, data sources, and research themes of studies using online user-generated text to discuss the medicinal use of cannabis?

## Materials and methods

### Search strategy

For this scoping review we used the PRISMA statement for systematic reviews and meta-analyses, to inform our systematic search methods (38).

Literature database queries were developed for four categories of studies. The first three categories used social media text as a data source, the fourth relied on internet search engine query data. For the first category, the database queries combined words used to describe social media forums, and cannabis-related keywords and general medical-related keywords (Table 1 Category 1). The second category also included the social media and cannabis-related keywords, but used keywords specific to psychiatric disorders, for which the use of medical cannabis has been described. Our search terms for this second category were informed by a systematic review of medicinal cannabis for psychiatric disorders (16) (Table 1 Category 2). The third category included social media and cannabis related keywords but focused on non-psychiatric medical conditions for which cannabis is sometimes used (Table 1 Category 3). The fourth category included studies using Internet search engine queries as a data source, there were no medical conditions included in these searches (Table 1 Category 4). A manual search of MEDLINE (Web of Science (1900-2022), Embase (OVID: 1974-2022), Web of Science Core Collection (1900-2022), and Scopus (1996-2022) databases was conducted by SKH and CMH in May 2021 and again in October 2022. The search was limited to English-language studies that were published between January 1974 and April 2022 (Appendix I).

**Table 1.**
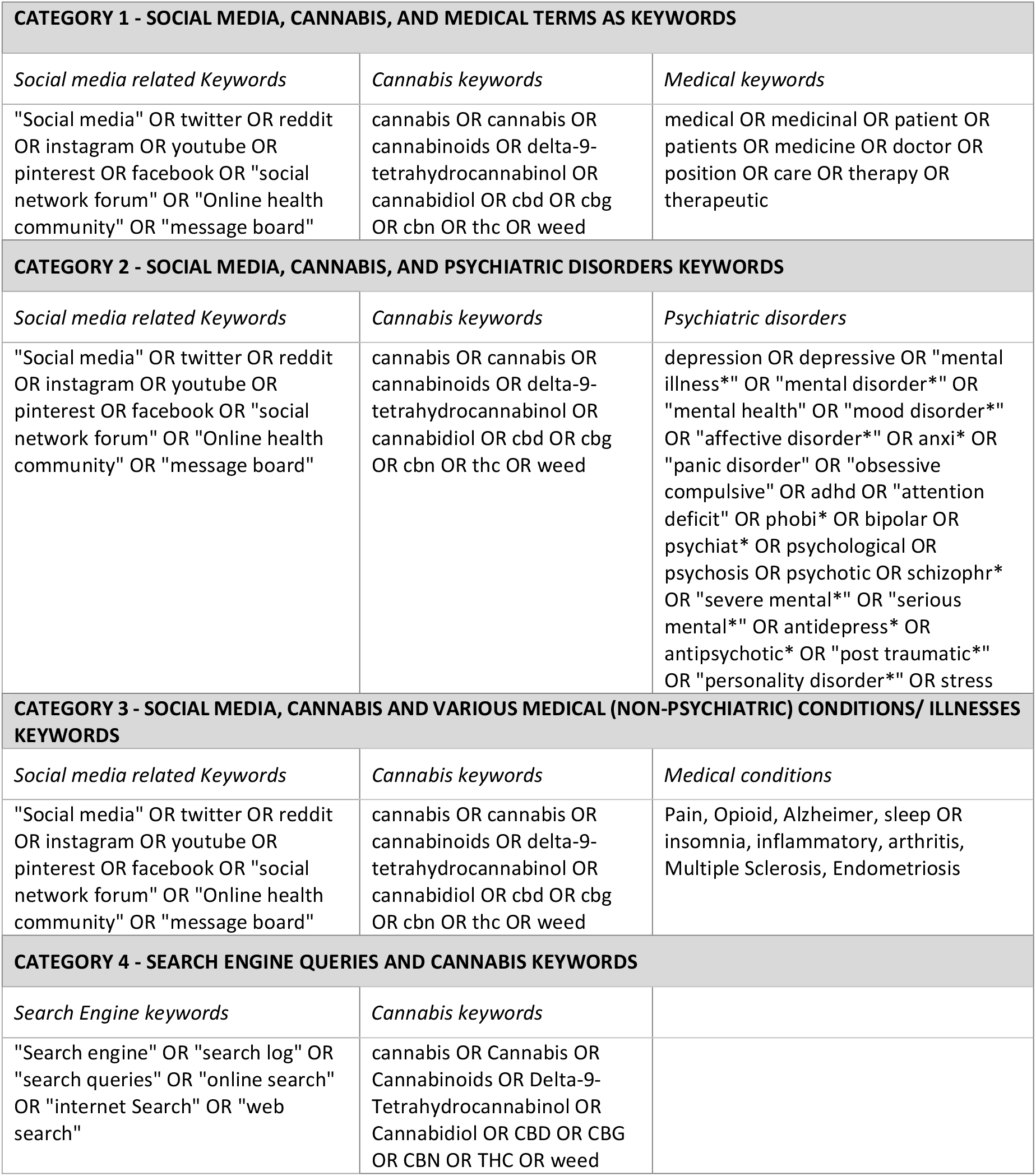
Search Strategy.

| **CATEGORY 1 - SOCIAL MEDIA, CANNABIS, AND MEDICAL TERMS AS KEYWORDS** |  |  |
| --- | --- | --- |
| *Social media related Keywords* | *Cannabis keywords* | *Medical keywords* |
| "Social media" OR twitter OR reddit OR instagram OR youtube OR pinterest OR facebook OR "social network forum" OR "Online health community" OR "message board" | cannabis OR cannabis OR cannabinoids OR delta-9-tetrahydrocannabinol OR cannabidiol OR cbd OR cbg OR cbn OR thc OR weed | medical OR medicinal OR patient OR patients OR medicine OR doctor OR position OR care OR therapy OR therapeutic |
| **CATEGORY 2 - SOCIAL MEDIA, CANNABIS, AND PSYCHIATRIC DISORDERS KEYWORDS** |  |  |
| *Social media related Keywords* | *Cannabis keywords* | *Psychiatric disorders* |
| "Social media" OR twitter OR reddit OR instagram OR youtube OR pinterest OR facebook OR "social network forum" OR "Online health community" OR "message board" | cannabis OR cannabis OR cannabinoids OR delta-9-tetrahydrocannabinol OR cannabidiol OR cbd OR cbg OR cbn OR thc OR weed | depression OR depressive OR "mental illness*" OR "mental disorder*" OR "mental health" OR "mood disorder*" OR "affective disorder*" OR anxi* OR "panic disorder" OR "obsessive compulsive" OR adhd OR "attention deficit" OR phobi* OR bipolar OR psychiat* OR psychological OR psychosis OR psychotic OR schizophr* OR "severe mental*" OR "serious mental*" OR antidepress* OR antipsychotic* OR "post traumatic*" OR "personality disorder*" OR stress |
| **CATEGORY 3 - SOCIAL MEDIA, CANNABIS AND VARIOUS MEDICAL (NON-PSYCHIATRIC) CONDITIONS/ ILLNESSES KEYWORDS** |  |  |
| *Social media related Keywords* | *Cannabis keywords* | *Medical conditions* |
| "Social media" OR twitter OR reddit OR instagram OR youtube OR pinterest OR facebook OR "social network forum" OR "Online health community" OR "message board" | cannabis OR cannabis OR cannabinoids OR delta-9-tetrahydrocannabinol OR cannabidiol OR cbd OR cbg OR cbn OR thc OR weed | Pain, Opioid, Alzheimer, sleep OR insomnia, inflammatory, arthritis, Multiple Sclerosis, Endometriosis |
| **CATEGORY 4 - SEARCH ENGINE QUERIES AND CANNABIS KEYWORDS** |  |  |
| *Search Engine keywords* | *Cannabis keywords* |  |
| "Search engine" OR "search log" OR "search queries" OR "online search" OR "internet Search" OR "web search" | cannabis OR Cannabis OR Cannabinoids OR Delta-9-Tetrahydrocannabinol OR Cannabidiol OR CBD OR CBG OR CBN OR THC OR weed |  |

The inclusion criteria for this review were: (i) peer reviewed research studies, (ii) peer reviewed conference papers (iii) studies which used online user-generated text as a data source, and (iv) social media research that was either directly focused on cannabis and cannabis products that have an impact on health or were health-related studies that found medicinal use of cannabis.

Exclusion criteria comprised: (i) editorials, letters, commentaries, surveys, protocols and book chapters; (ii) studies that used social media for recruiting participants; (iii) studies where the full text of the publication was not available; (iv) conference abstracts (iv) studies primarily focused on electronic nicotine delivery systems adapted to deliver cannabinoids; (v) studies that used bots or autonomous systems as the main data source and (vi) studies that focused exclusively on synthetic cannabis.

All studies captured by the search queries listed in Table 1 were uploaded into excel to enable all duplicates to be removed. Following this all titles and abstracts were reviewed independently and in duplicate by CMH and SKH. Records were excluded based on title and abstract screening as well as publication type.

The full text articles that were identified for inclusion following screening process were then independently critiqued by pairs of reviewers using a checklist developed for this study. The purpose of the checklist that we developed for this systematic scoping review was to provide an overall assessment of quality rather than generate a specific score, (Appendix 1). Assessments of quality in each study were based on evidence of relative quality in the aims or objectives, main findings, data collection method, analytic methods, data source, and evaluation and interpretations of the study. CMH and SKH critiqued all articles, and YB and MC each critiqued a selection of studies to ensure each article had been independently reviewed by two researchers. Where initial disagreement existed between reviewers regarding the inclusion of a study, team members met to discuss the disputed article’s status until consensus was achieved.

### Study inclusion

Assessments of quality in each study were based on evaluating each study’s aims and objectives, main findings, data collection and analytic methods, data sources, and evaluation and interpretations of the results. Social media studies were included if there were no major biases affecting the internal, external or construct validity of the study (39). In doing so, the internal validity of each study was determined by the quality of the data and analytic processes used, the external validity determined by the extent to which the findings can be generalized to other contexts, and the construct validity was ascertained by the extent to which the chosen measurement tool correctly measured what the study aimed to measure (Appendix II).

Of the 1,556 titles identified in the electronic database searches, 858 duplicate articles were removed, 449 were excluded following the screening of title and abstracts and 197 were excluded based on publication type (i.e., survey, letter, comment, abstract). This screening process provided 52 potentially relevant full text primary research studies to be included in the review. Of these, five articles were not able to be retrieved, two out of 47 articles had initial disagreement. Upon consensus, five were excluded with reasons (Appendix III) using the quality assessment checklist as described above. This provided 42 papers for inclusion in this systematic scoping review.

The PRISMA flow diagram is presented in Fig 1.

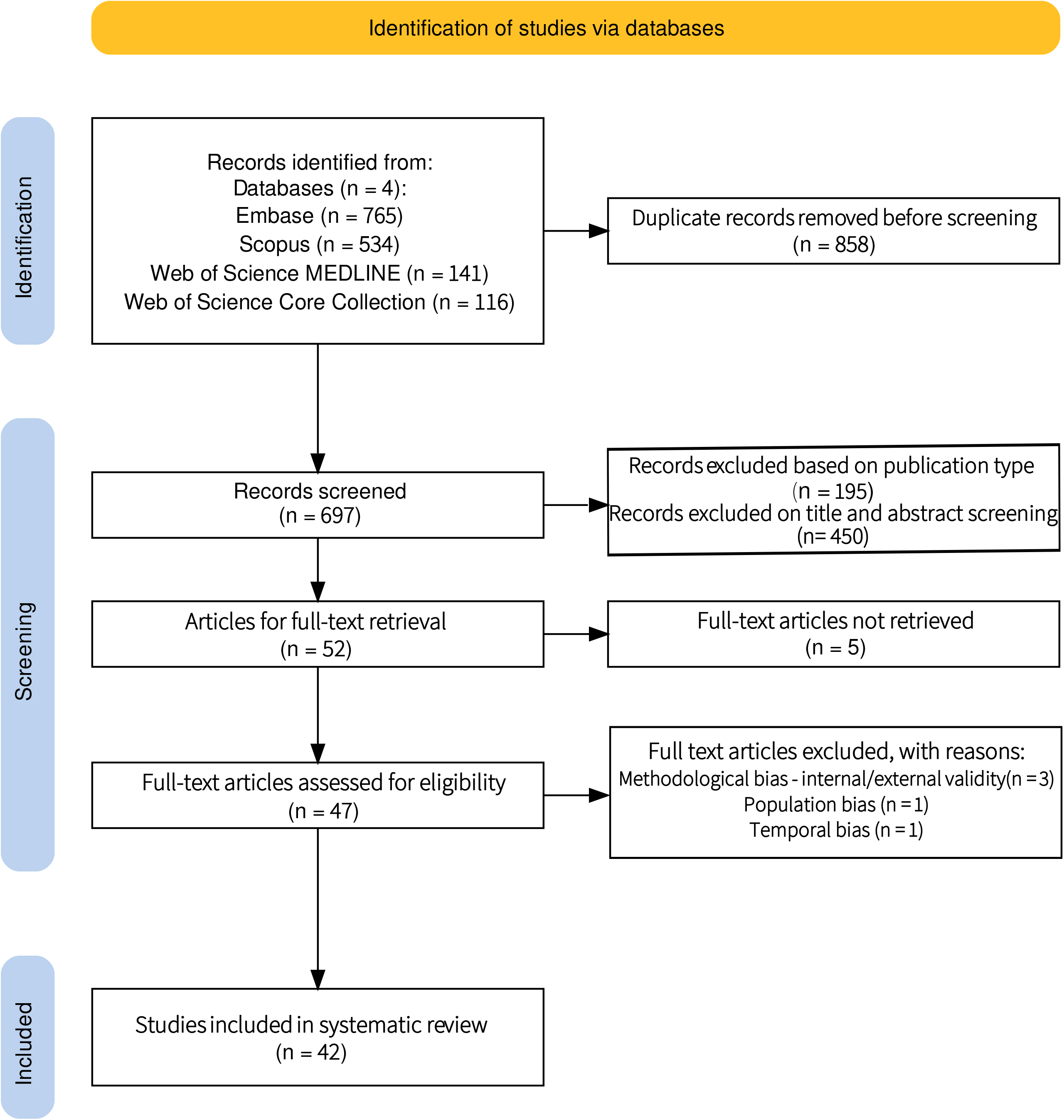

## Results

Of the 42 papers published between 2014 and 2022 included in this systematic scoping review, most were journal articles 40/42 (95.2%), and two were conference-based publications 2/42 (4.8%). Although the first study was published eight years ago, nearly two-thirds 24/42 (57%), have been published over the last four years. Table 2 provides a summary of each paper that includes author names, publication year, data source, duration of the study, number of collected posts, number of analyzed posts, and the coding/labelling approach used.

**Table 2.**
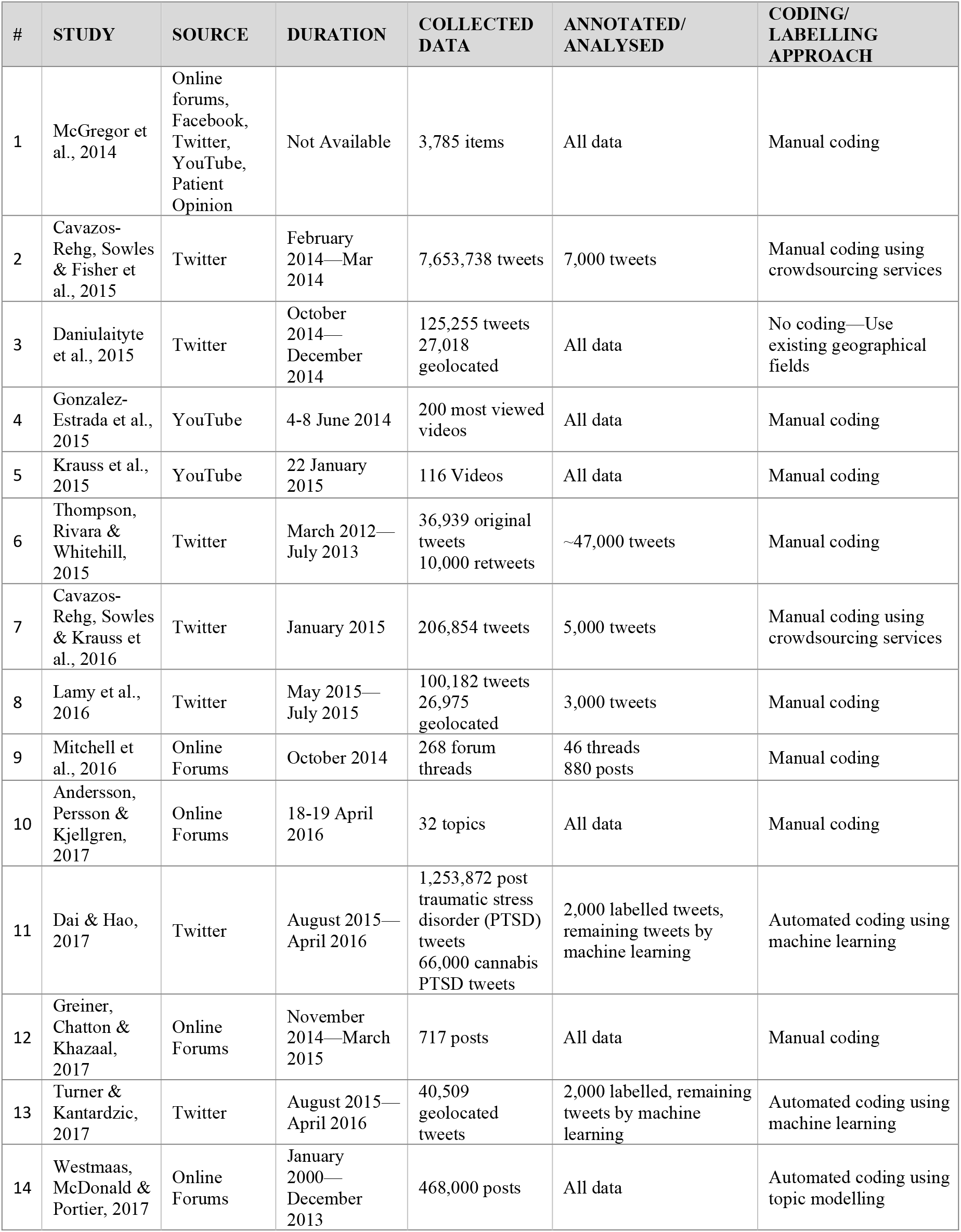
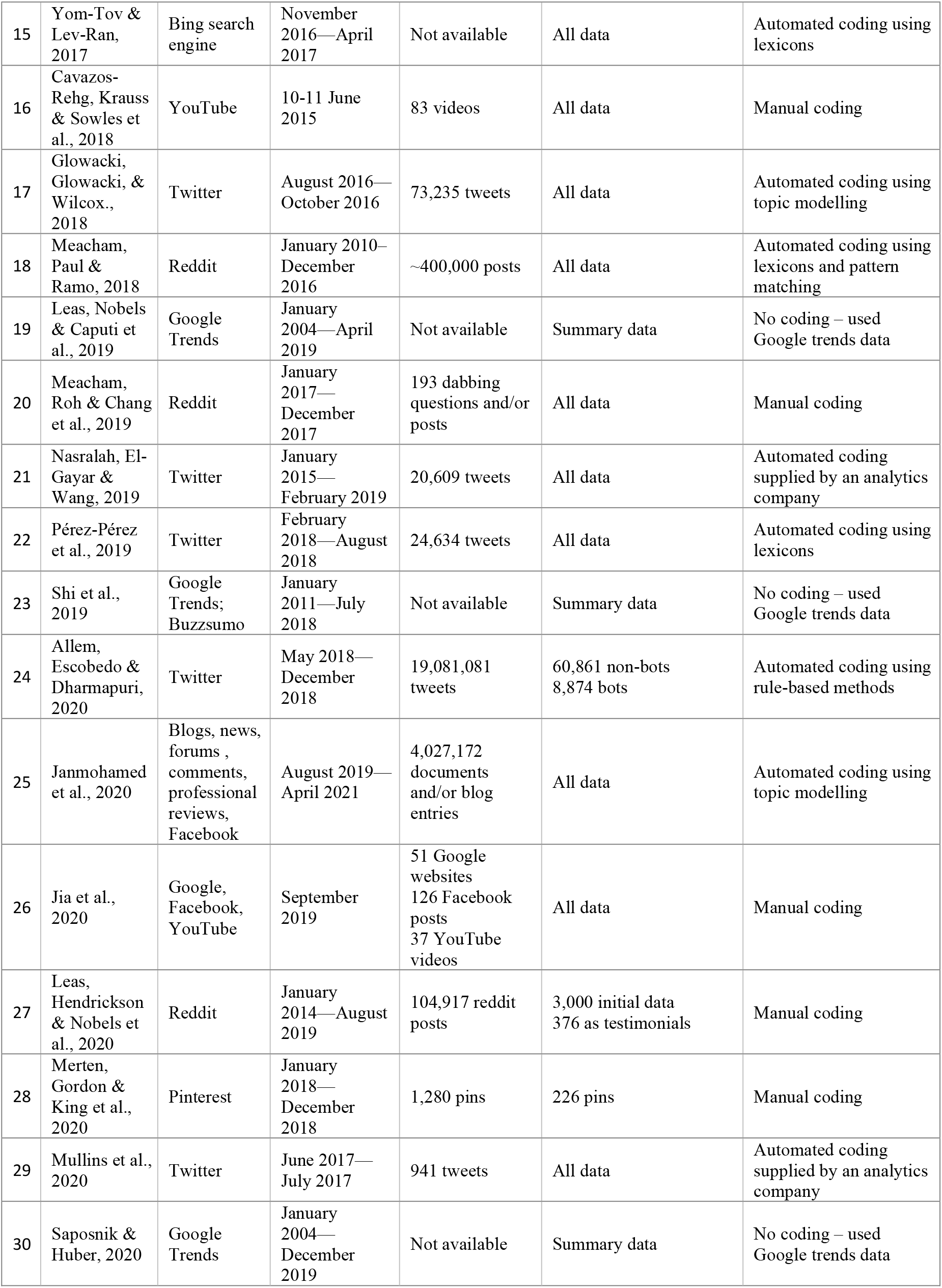
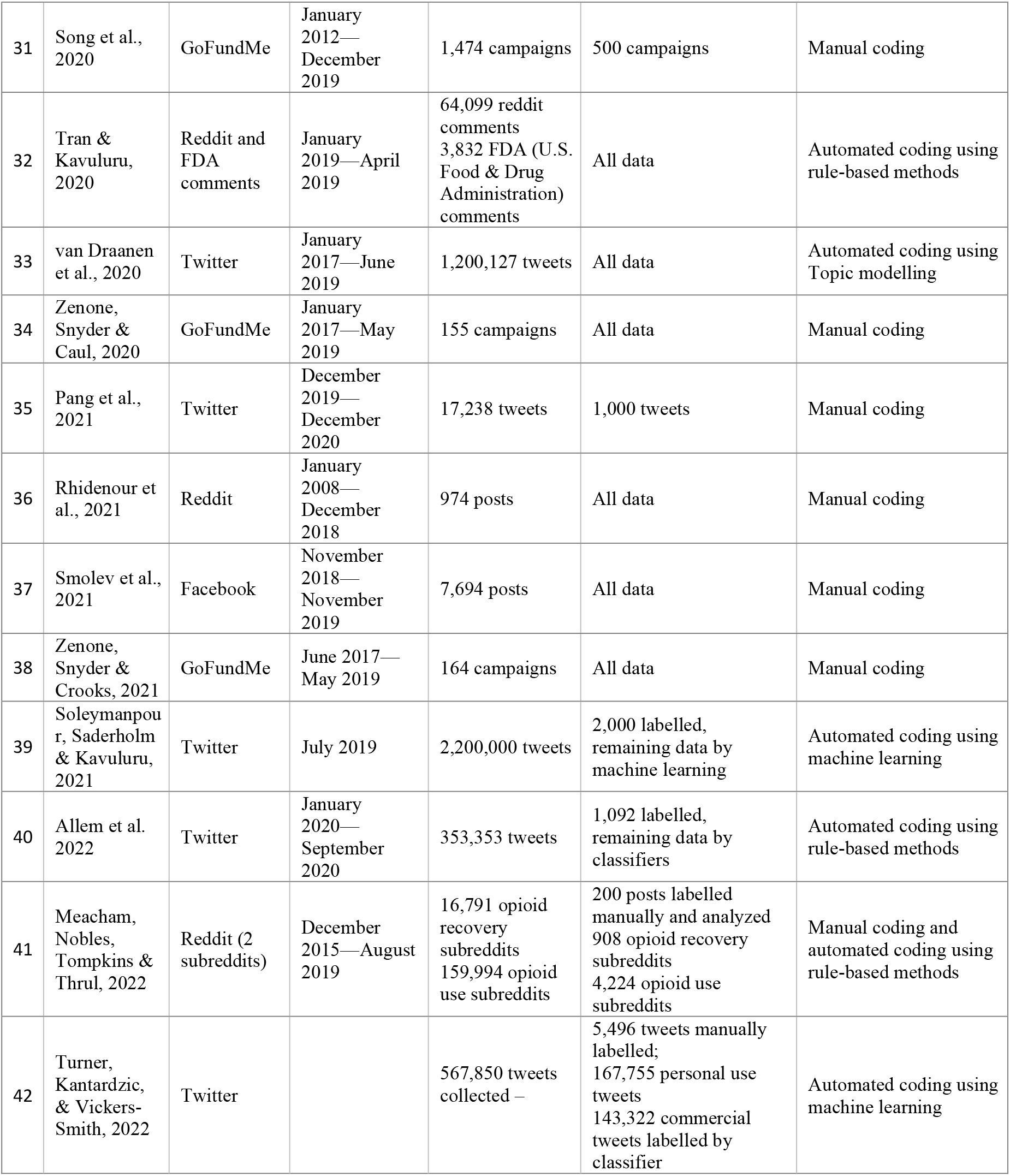
Articles included in the review.

### Data collection and annotation

The largest manually annotated dataset that contained 47,000 labelled tweets was published by (40) in 2015. This paper was one of 22 studies included in this review (54.8%) that either collected a limited number of data points, or sampled their collected data, and manually coded the data to gain an in-depth understanding of the domain (41-62). Four of the 42 studies (9.5%) used existing meta-data including Google trends summary data (63-65), and geo-location data (66). Two (4.8%) studies used data that was manually coded using crowdsourcing services (42, 46), and two (4.8%) used automated coding supplied by a social media analytics company (67, 68),. Fifteen of the 42 studies (35.7%) used automated methods for labelling data, that included the use of machine learning, lexicon, and rule-based algorithms (57, 69-82). Automated coding was increasingly used as an analytic tool for social media data on this topic from 2017 onward (Table 2).

### Data analysis

The manually labelled studies calculated proportions and trends and developed themes (40, 41, 43, 44, 46, 47, 51-53, 57, 58, 61, 62, 79, 83-91). (63-65) reported on Google trends data that delivered an index of Google search trends over time. (66) processed Twitter data that contained existing geographical fields to identify geolocation at source. The studies that utilised a large volume of data used advanced computational methods, which included sentiment analysis, topic modeling, and rule-based text mining (57, 69, 74-79, 82). The use of sentiment analysis in the (57, 77, 92) enabled the analysis of people’s sentiments, opinions, and attitudes. Topic modelling in the (71, 73, 76, 77, 82) studies enabled the development of themes via automatic machine learning methods. The use of rule-based text mining such as found in the (57, 69, 80, 93, 94) studies enabled the classification of posts into pre-existing health-related categories.

### Research Themes

In this review, we categorized the 42 research articles into six broad themes. Themes were based on the research questions motivating the studies, where each paper was classified as belonging a primary theme, based on alignment with the research aims.

### General cannabis-related

Nine studies were included in this theme (Table 3). The main keywords used in these studies included general terms such as ‘cannabis’, ‘marijuana’, ‘pot’ and ‘weed’. The major aim of these studies was to either identify topics of conversations regarding cannabis, or to examine their sentiments. These studies are included because they reported on conversations around cannabis use for medical purposes, sentiment associated with perceptions of health benefits of cannabis, and reports of adverse effects. For example, a study on veterans use of cannabis found that cannabis is used to self-medicate a number of health issues, including Post-Traumatic Stress Disorder (PTSD), anxiety and sleep disorder (91). Seven of the studies used Twitter as a data source (40, 70, 75, 84, 93, 95, 96), one examined the content of YouTube videos about cannabis (97), one investigated online self-help forums (85) and another used Reddit data (91).

**Table 3.** Papers studying general conversations.

| STUDY | AIM | HEALTH-RELATED EFFECTS/CLAIMS | DATA IDENTIFICATION |
| --- | --- | --- | --- |
| Cavazos-Rehg, Krauss et al., 2015 | To examine the sentiment and themes of cannabis-related tweets from influential users and to describe the users' demographics. | A common theme of pro tweets was that cannabis has health benefits. Anti-cannabis posts spoke of the harm experienced in using cannabis. 77% of posts had positive sentiments, with 12 times higher reach than other posts. | Cannabis-related keyword |
| Thompson, Rivara et al., 2015 | To examine cannabis- related content in Twitter, especially content tweeted by adolescent users, and to examine any differences in message content before and after the legalization of recreational cannabis in two US states. | More tweets described perceived positive benefits of cannabis use, including relaxation and escaping life problems. Tweets described cannabis as less harmful than other drugs or as not harmful at all and suggested its medical role for conditions such as depression and cancer. Less than 1% of tweets expressed a concern about cannabis use. | Cannabis-related keyword |
| Greiner, Chatton et al., 2017 | To investigate online content of cannabis use/addiction self-help forums. | Self-help forums on cannabis share a theme around cannabis users seeking help for addiction and withdrawal issues. | keywords "cannabis" and "forum" and "help" |
| Turner et al., 2017 | To examine if cannabis legalization policies impact Twitter conversations and the social networks of users contributing to cannabis conversations. | Medical cannabis was a major topic in the conversations. | Cannabis-related keywords |
| Cavazos-Rehg, Krauss et al., 2018 | To investigate cannabis product reviews and the relationship between exposure to product reviews and cannabis users' demographics and characteristics. | Product reviews promoted cannabis for helping with relaxation, pain relief, sleep, improving emotional well-being. Medical cannabis users are more likely to be exposed to cannabis product reviews. | "weed review", "marijuana review", "cannabis review" |
| Allem, Escobedo et al., 2020 | To identify and describe cannabis-related topics of conversation on Twitter, and the public health implications of these. | Health and medical was the third most prevalent topic of the 12 topics identified in the data. Posts suggested that cannabis could help with cancer, sleep, pain, anxiety, depression, trauma, and posttraumatic stress disorder. Health-related posts from social bots were almost double of genuine posts. | Cannabis-related keywords |
| Van Draanen et al., 2020 | To examine differences in the sentiment and content of cannabis-related tweets in the US (by state cannabis laws) and Canada. | Medical cannabis use was one of the main topics of conversations in cannabis-related tweets from both countries. | Tweets filtered on US and Canada geolocation and then further filtered on cannabis-related keywords |
| Rhidenour et al., 2021 | To explore Veterans' Reddit discussions regarding their cannabis use. | Over a third of the Reddit posts described the use of medical cannabis as an aid for psychological and physical ailments. Overall, veterans discussed how the use of medical cannabis reduced PTSD symptoms, anxiety, and helped with their sleep. | The veteran subreddit |
| Allem, Majmundar et al., 2022 | To determine the extent to which a medical dictionary could identify cannabis-related motivations for use and health consequences of cannabis use. | There were posts related to both health motivations and consequences of cannabis use. The health-related posts included issues with the respiratory system, stress to the immune system, and gastrointestinal issues, among others. | Cannabis-related keywords |

### Cannabis mode of use

Seven studies reported on the mode of use of cannabis as a medicine (Table 4). These studies collected data using keywords such as ‘vape’, ‘vaping’, ‘dabbing’, and ‘edibles’. Conversations around modes of use revealed a theme about lacking, seeking, or sharing knowledge about possible health consequences of the modes of use. Another theme was around the perceived health benefits of cannabis and the various modes of use of cannabinoids that included sleep improvement and relaxation resulting from dabbing oils (46) or consuming ‘edibles’ (47). The findings suggest that for emerging modes of use such as dabbing, where the availability of evidence-based information is limited, people seek information from others’ experiences.

**Table 4.** Papers studying Cannabis mode of use.

| STUDY | AIM | HEALTH-RELATED EFFECTS/CLAIMS | DATA IDENTIFICATION |
| --- | --- | --- | --- |
| Daniulaityte et al., 2015 | To explore Twitter data on concentrate ('dabs') use and examine the impact of cannabis legalization policies on concentrate use conversations. | Twitter data suggest popularity of dabs in the US states with legalized recreational/medical use of cannabis. Dabbing as an emerging mode of use could carry significant health risks. | dab-related keywords for U.S. location |
| Krauss, Sowles et al., 2015 | To explore the content of cannabis dabbing-related videos on YouTube. | Only 21% of videos contained some warning about dabbing, such as preventing explosions, injury, or negative side effects. 22% of videos specifically mentioned medical cannabis or getting "medicated", either in the video itself or in the accompanying text description. | Dabbing related keywords |
| Cavazos-Rehg, Sowles et al., 2016 | To study themes of dabbing conversations and to investigate the consequences of high-potency cannabis consumption. | 4th theme (of 7 themes) was about cannabis helping with relaxation, sleep or solving problems. Extreme effects were both physiological and psychological. The most common physiologic effects were passing out and respiratory, with coughing the most common respiratory effect. | Dabbing related keywords |
| Lamy, Daniulaite et al., 2016 | To study themes of edibles conversations and examine legalization policies' impact on cannabis-related tweeting activity. | Twitter data suggest mostly positive attitudes toward cannabis edibles. Positive tweets describe the quality of the "high" experienced and how cannabis edibles facilitate falling asleep. Negative tweets discuss the unreliability of edibles' THC dosage and delayed effects that were linked to over-consumption, which could lead to potential harmful consequences. | Cannabis edible-related keywords |
| Meacham, Paul et al., 2018 | To analyse discussions of emerging and traditional forms of cannabis use. | Less than 2% of conversations described adverse effects. The most mentioned adverse effects were anxiety-related in the context of smoking, edibles, and butane hash oil, and cough for vaping and dabbing. | A cannabis specific subreddit on various modes of use |
| Meacham, Roh et al., 2019 | To study themes of dabbing-related questions and responses. | Health concerns are the 5th category of dabbing questions - including respiratory effects, anxiety, and vomiting. | Search for "Dab" and "question" |
|  |  | Respondents in these conversations usually spoke from personal experience. | on cannabis subreddits |
| Janmohamed et al., 2020 | To map temporal trends in the web-based vaping narrative, to indicate how the narrative changed from before to during the COVID-19 pandemic. | The emergence of a vape-administered CBD treatment narrative around the COVID-19 pandemic. | Vape-related keywords |

### Cannabis as a medicine for a specific health issue

Six studies were included in this theme (Table 5). These studies investigated conversations around the use of cannabis or cannabidiol for a specific health issue. The health conditions included glaucoma (53), PTSD (69), cancer (62, 98), Attention Deficit Hyperactivity Disorder (ADHD) (88) and pregnancy (89). These studies mostly discovered that conversations claimed benefits of cannabis as an alternative treatment for these health conditions, although mentions of harm, and both harm and therapeutic effects, were also present (88).

**Table 5.** Papers studying Cannabis as a medicine for a specific health issue.

| STUDY | AIM | HEALTH-RELATED EFFECTS/CLAIMS | DATA IDENTIFICATION |
| --- | --- | --- | --- |
| Mitchell et al., 2016 | To examine the content of online forum threads on ADHD and cannabis use to identify trends about their relation, particularly regarding therapeutic and adverse effects of cannabis on ADHD. | 25% of individual posts indicated that cannabis is therapeutic for ADHD, as opposed to 8% that claim it is harmful, 5% that it is both therapeutic and harmful, and 2% that it has no effect on ADHD. | Cannabis and ADHD keywords |
| Dai & Hao, 2017 | To evaluate factors that could impact public attitudes to PTSD related cannabis use. | 5.3% of all PTSD tweets were related to cannabis use and these tweets predominantly supported cannabis use for PTSD. | Cannabis and PTSD keywords |
| Shi et al., 2019 | To characterize trends in use of cannabis for cancer and analysis of content and impact of popular news about cannabis for cancer. | Between 2011-2018, the relative google search volume of 'cannabis cancer' queries increased at a rate 10 times faster than 'standard cancer therapies' queries. Popular 'false news' stories had a much higher engagement than contrary 'accurate' news stories. | Cannabis vs standard therapies for cancer |
| Jia, Mehran et al., 2020 | To analyze the content quality and risk of readily available online information regarding cannabis and glaucoma. | Although the American Glaucoma Society recommends against cannabis use for glaucoma treatment, 21% of Facebook, 24% of Google, and 59% of YouTube search results were pro cannabis use for glaucoma treatment. | Cannabis and Glaucoma keywords |
| Zenone et al., 2020 | To use crowdfunding campaigns to understand how cannabidiol is represented/misrepresented as a cancer-related care. | CBD use was reported to reduce the side effects of conventional treatments or can be used with other complementary cancer treatments. Reported uses included stimulating appetite, general pain relief, assisting with sleep, countering nausea, or general recovery purposes. Most campaigners presented definite efficacy of CBD for pain or symptom management. | CBD term variants and "cancer" |
| Pang et al., 2021 | To examine cannabis and pregnancy-related tweets over a 12-month period. | 36% mentioned safety during pregnancy, 2.3% of posts asked about safety during postpartum, and 2.7% of posts expressed use of cannabis during pregnancy to help with pregnancy symptom i.e. to help with morning sickness, nausea, vomiting, headaches, pain, stress, and fatigue. The authors conclude that health providers discuss risks and provide official information about cannabis use in pregnancy. | Cannabis and pregnancy related keywords |

### Cannabis as a medicine as part of discourse on illness and disease

Eleven studies were included in this theme. In this category, the research focus was on social media topics relating to management and treatment options for a range of health conditions rather than on medicinal cannabis per se (Table 6). Health conditions included inflammatory and irritable bowel disease(99), opioid use disorder (67, 100, 101), pain (68), ophthalmic disease (3), cluster headache and migraine(83), asthma (102), cancer (90, 103), autism disorder (104) and brachial plexus injury (61).

**Table 6.**
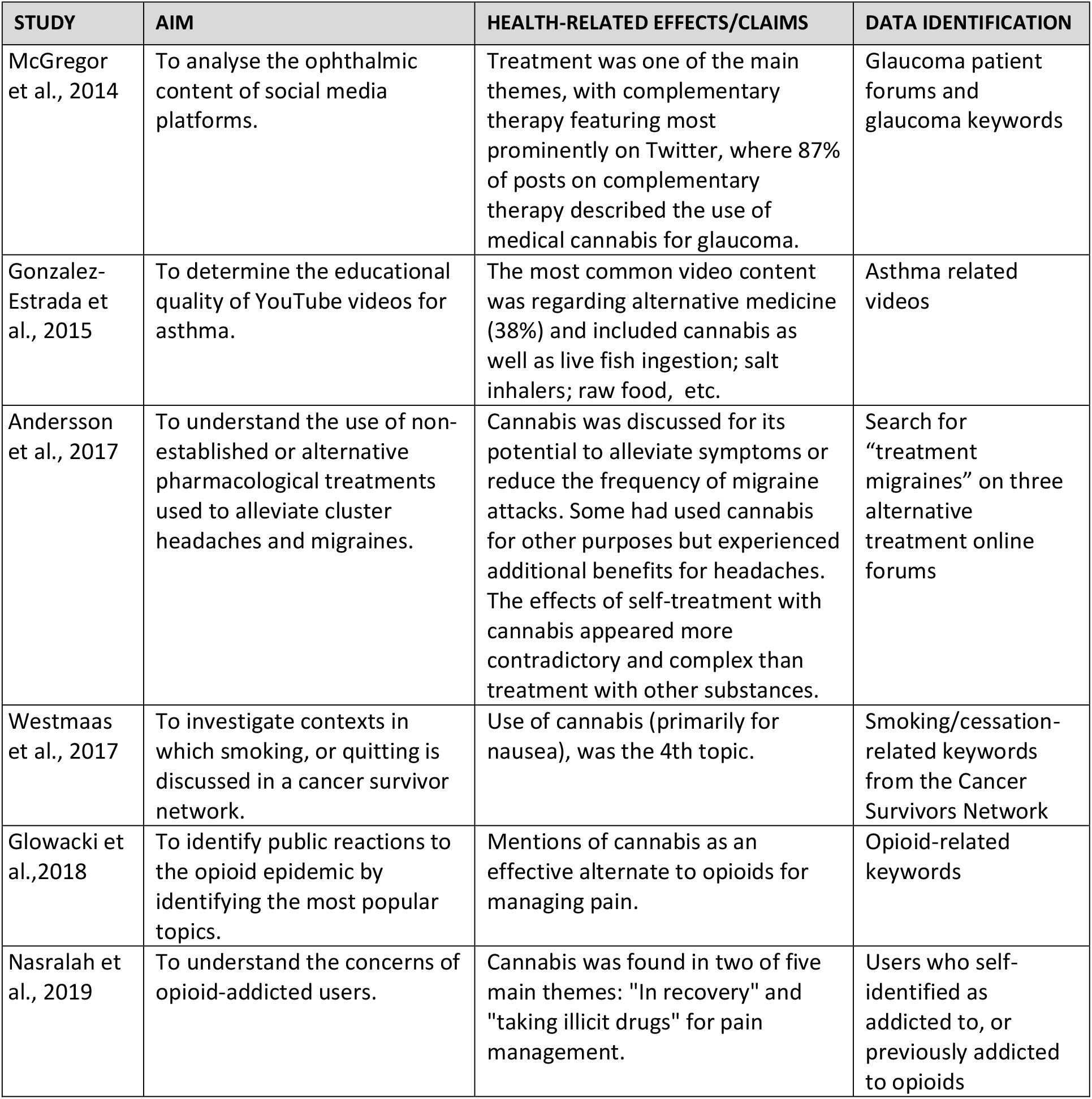
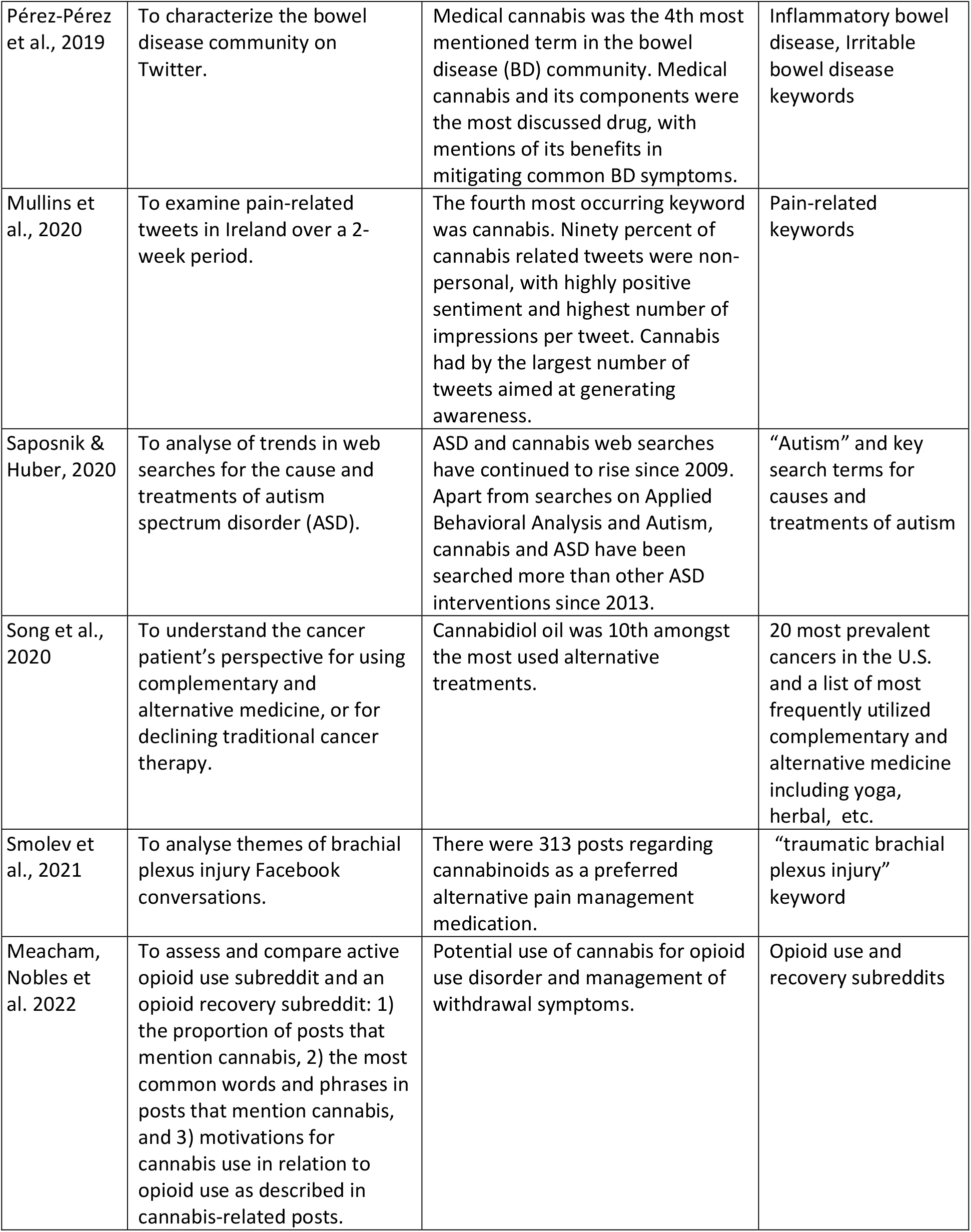
Papers studying Cannabis as a medicine as part of discourse on illness and disease.

### Cannabidiol (CBD)

There were six studies in the cannabidiol (CBD) category (58, 77, 79, 86, 87, 105, 106) (Table 7). These studies concentrated on conversations related to the benefits of CBD products, product sentiment (positive, negative, or neutral), the factors that impact on a person’s decision to use CBD products, and the trends in therapeutic use of CBD.

**Table 7.** Papers studying Cannabidiol (CBD)

| STUDY | AIM | HEALTH-RELATED EFFECTS/CLAIMS | DATA IDENTIFICATION |
| --- | --- | --- | --- |
| Merten, Gordon et al., 2020 | To analyse how CBD is portrayed on Pinterest. | Most pins (57.5%) did not make a specific health benefit claim yet 42.5% claimed mental, physical, or both mental and physical health benefits. | 'cannabidiol' or 'CBD' |
| Zenone, Snyder et al., 2021 | To analyse the CBD informational pathways which bring consumers to CBD for medical purposes. | Self-directed research was the most common pathway to CBD. The proposed uses of CBD were for cancer, seizure-inducing diseases/conditions, joint/inflammatory diseases, mental health disorders, nervous system diseases, and autoimmune diseases. | 'cannabidiol' or 'CBD' |
| Leas, Nobel et al., 2019 | To analyse public interest trends in CBD using Google Trends. | Searches for CBD exceed searches for yoga and around half as much as searches for dieting. | 'cannabidiol' or 'CBD' vs other alternative medicine including diet, yoga, etc. |
| Leas, Hendrickson et al., 2020 | To assess if individuals are using CBD for diagnosable conditions which have evidence-based therapies. | Psychiatric conditions were the most cited diagnosable condition, mentioned in 63.9% of testimonials. The second most cited subcategory was orthopedic conditions (26.4%), followed by sleep (14.6%), neurological (6.9%), and gastroenterological (3.9%) conditions. | CBD subreddit posts |
| Tran & Kavuluru, 2020 | To examine social media data to determine perceived remedial effects and usage patterns for CBD. | Anxiety disorders and pain were the two conditions dominating the discussion surrounding CBD, both in terms of general discussion and for CBD as a perceived therapeutic treatment. CBD is mentioned as a treatment for mental issues (anxiety, depression, stress) and physiological issues (pain, inflammation, and others) | CBD subreddit posts |
| Soleymanpour et al., 2021 | To perform content analysis of marketing claims for CBD in Twitter. | Over 50% of CBD tweets appear to be marketing related chatter. Pain and anxiety are the most popular conditions mentioned in marketing messages. Edibles are the most popular product type being advertised, followed by oils. | "cbd", "cbdoil" and "cannabidiol" |
| Turner, Kantardzic et al. 2021 | The objective of this study was to provide a framework for public health and medical researchers to use for identifying and analyzing the consumption and marketing of unregulated substances using CBD as an exemplar. | There was a significant difference in the sentiment scores between the personal and commercial CBD tweets, the mean sentiment score of the commercial CBD scores was higher than that of the personal CBD score. Pain, anxiety, and sleep had the highest positive sentiment score for both personal and commercial CBD tweets. | "CBD", "cannabidiol." |

### Adverse drug reactions and adverse effects

One paper had a research question that explicitly focused on the detection of adverse events (72) (Table 8) This study explored the prevalence of internet search engine queries relating to the topic of adverse reactions and cannabis use. Seven other studies contained mentions of adverse effects which were associated with cannabis use (44, 46, 48, 50, 52, 59, 74, 78), however these papers were not included under this theme, as their research questions were not centred around the explicit investigation of adverse events.

**Table 8.** Papers studying adverse events related to cannabis.

| STUDY | AIM | HEALTH-RELATED EFFECTS/CLAIMS | DATA IDENTIFICATION |
| --- | --- | --- | --- |
| Yom-Tov & Lev-Ran, 2017 | To check if search engine queries can be used to detect adverse reactions of cannabis use. | A high correlation between the side effects recorded on established reporting systems and those found in the search engine queries. These side effects included anxiety, depression-related symptoms, psychotic symptoms such as paranoia and hallucinations, cough and other symptoms. | Cannabis related keywords |

## Discussion

Currently, there exist systematic reviews of cannabis and cannabinoids for medical use based on clinical efficacy outcomes from randomized clinical trials (18) and reviews on the use of social media for illicit drug surveillance (107). However, following searches on PROSPERO and the databases listed above, to our knowledge, this paper constitutes the first systematic scoping review examining studies that used user-generated online text to understand the use of cannabis as a medicine in the global community.

Our scoping review found that the use of social media and internet search queries to investigate cannabis as a medicine is a rapidly emerging area of research. Over half of the studies included in this review were published within the last three years, this reflects not only increase community interest in the therapeutic potential of cannabinoids, but also world-wide trends towards cannabis legalization (4, 7-9, 108, 109). Regarding social media platforms, Twitter was the data source in eighteen (42.9%) of the 42 studies, almost three and a half times the number of studies using Reddit (6, 14.3%) and just under three times the number of studies using data from Online forums (5, 11.9%). Three (7.14%) GoFundMe studies and three (7.14%) Google Trends studies were also included in the review. Hence, much of the data in this systematic review comprised posts from the Twitter platform. Several factors may explain this finding, firstly Twitter is real-time in nature, it has a high volume of messages, and it is publicly accessible. These factors makes it a useful data source for public health surveillance (110).

Regarding the subjects of the studies, twelve (28.6%) focused on general user-generated content regarding the treatment of health conditions (glaucoma, autism, asthma, cancer, bowel disease, brachial plexus injury, cluster headaches, opioid disorder). These studies were either explicitly designed to investigate cannabis as a medicine or were studies that generated results that incidentally found cannabis mentioned as an alternative or complimentary treatment (either formally prescribed or via self-medication).

Qualitative studies featured in the research, but while their contributions are valuable, especially in the context of hypothesis generation, they tend to be limited by their smaller datasets, which frequently comprised manually annotated samples. The recent emergence of powerful machine learning-based natural language processing (NLP) models suggests that it should be possible to automate the continuous processing of far larger datasets using NLP technologies, built upon the insights gained from initial qualitative studies, and even leveraging their annotated data for training purposes. Recent trends in the social science data landscape have shown a convergence between social science and computer science expertise, where the ability to use computational methods has greatly assisted the collection and validation of robust datasets that can form the basis of deeper social science research (111).

We found much heterogeneity in approaches applied to analyse user-related content, and inconsistent quality in the methodologies adopted. While we endeavored to include as many studies as possible, some of the publications initially identified as suitable for inclusion were not suitable based on a minimum quality requirements checklist (Appendix 1). This checklist was designed to ensure that selection of data source, choice of platform, data acquisition and preparation, analysis and evaluation delivers data and conclusions that are appropriate for answering the research questions.

The utilization of user-generated content for health research is subject to several inherent limitations which include the lack of control that researchers have in relation to the credibility of information, the frequently unknown demographic characteristics and geographical location of individuals generating content, and the fact that social media users are not necessarily representative of the wider community (112). Furthermore, the uniqueness, volume, and salience of social media data has implications that need to be considered when used for health information analysis (113). Volume is usually inversely related to salience: a platform such as Twitter has a very high volume of information, but, apart from its frequency, much of which is not highly pertinent for analyzing an effect; whereas the volume of information contained in a blog will much less but likely more salient for analysis. Notwithstanding these limitations, user-generated content comprises large-scale data that provides access to the unprompted organic opinions and attitudes of cannabis users in their own words and is an effective medium through which to gauge public sentiment. To date, insights regarding cannabis as medicine have gained primarily through surveys or focus groups which have their own limitations regarding the format of data collection and potential bias in participant recruitment. A limitation of this scoping review was the lack of inclusion of a computational database such as *IEEE Xplore* in the search strategy, and the exclusion of the search terms ‘infodemiology’ and ‘infoveillance’. Infodemiology and infoveillance studies explicitly use web-based data for research, and *IEEE Xplore* is a repository that contains technical papers and documents relating to computer science. However, our search was systematic, comprehensive and *IEEE Xplore* is Scopus-indexed, and we expect data loss to be minimal.

## Conclusion

Our systematic scoping review reflects a growing interest in the use of user-generated content for public health surveillance. It also demonstrates there is a need for the development of a systematic approach for evaluating the quality of social media studies and highlights the utility of automatic processing and computational methods (machine learning technologies) for large social media datasets. This systematic scoping review has shown that user-generated content as a data source for studying cannabis as a medicine provides another means to understand how cannabis is perceived and used in the community. As such, it is another potential ‘tool’ with which to engage in pharmacovigilance of, not only cannabis as a medicine, but also other novel therapeutics as they enter the market.

## Data Availability

N/A

## Acknowledgments

This work was supported by the Australian Centre for Cannabinoid Clinical and Research Excellence (ACRE), funded by the National Health and Medical Research Council (NHMRC) through the Centre of Research Excellence scheme (NHMRC CRE APP1135054).

## Supporting information

Appendix I. Search strategies for each database (Doc)

Appendix II. Quality assessment checklist (Doc)

Appendix III. Excluded studies (Doc)

S1. Database queries (Doc)

S2. Database queries results (Xlsx)

S3. PRISMA Checklist (Doc)

